# Sulfated N-glycans Upregulation in Sera Predicts Early-Stage Breast Cancer in Patients

**DOI:** 10.1101/2024.03.27.24305000

**Authors:** Dereje G. Feleke, Bryan M. Montalban, Solomon T. Gizaw, Hiroshi Hinou

**Affiliations:** Graduate School of Life Science and Faculty of Advanced Life Science, Frontier Research Center for Advanced Material and Life Science, Hokkaido University, N21, W11, Sapporo 001-0021, Japan; Department of Chemistry, College of Science, De La Salle University, 2401 Taft Avenue, Manila 0922, Philippines; Department of Biochemistry, College of Health Science, Addis Ababa University, Addis Ababa, Ethiopia

## Abstract

Alterations in sulfated glycans are associated with several pathological conditions, including cancer. However, analysis of sulfated glycans poses challenges, making the investigation of sulfated glycan profiles a topic of significant interest in the search for novel biomarkers for early BC detection. We used a glycoblotting-based sulphoglycomics workflow to examine sulfated N-glycans present in the serum of Ethiopian patients with BC. Seven mono-sulfated glycans were significantly upregulated in the sera of BC patients compared to the control group. Each identified glycan showed significant abundance with AUC ≥ 0.8 and demonstrated high diagnostic accuracy in predicting early stage BC patients. Importantly, the sulfated glycans were analyzed without removing the sialic group, allowing for comprehensive evaluation of the sialylation status of the identified sulfated glycans. This study represents the first quantitative analysis of sulfated N-glycans in patients with BC and identifies novel biomarkers with discriminatory potential in the early stages of BC.

**Statement of significance:** This study presents a quantitative analysis of sulfated N-glycans in BC, aiming to identify novel glyco-biomarkers that demonstrate high diagnostic accuracy for early stage BC. Analyzing sulfated glycans without removing sialic acids offers comprehensive insights. These findings advance the understanding of BC, potentially enhance early detection, and improve patient outcomes.

## Introduction

Breast cancer (BC) is the most prevalent cancer and the leading cause of cancer-related death among women worldwide. ^1^ Early detection and diagnosis are crucial for improving the survival rates of patients with BC. Currently, imaging methods, such as mammography, magnetic resonance imaging, and ultrasonography, are the primary methods for BC screening. However, these methods are invasive and time consuming. ^2, 3^ Serum biomarkers such as Cancer Antigen 15-3 (CA 15-3) and carcinoembryonic antigen (CEA) have also been explored. However, these biomarkers lack sensitivity and specificity and are used to monitor treatment responses in patients. ^4, 5^ Therefore, highly sensitive and specific biomarkers for early stage BC diagnosis are needed. Glycosylation, a common post-translational modification (PTM), plays crucial roles in various biological processes, including molecular recognition, adhesion, migration, immune regulation, and receptor signaling. ^6–8^ Altered glycosylation profiles of glycoproteins have been implicated in tumor cell proliferation, differentiation, invasion, and metastasis. ^9–11^ These changes correlate with the tumor burden and poor prognosis, making glycosylation analysis valuable for distinguishing between normal and cancerous cells. ^2, 7^ Previously, our group also explored altered non-sulfated glycans associated with the invasiveness and metastatic potential of BC. ^12^

Moreover, glycans can undergo modifications, such as sulfation, phosphorylation, and acetylation, with sulfation being the most common post-glycosylation modification. ^13^ Sulfated glycans are involved in physiological processes, such as lymphocyte homing^14^ and hormone clearance^15^, as well as in diseases, such as cystic fibrosis^16^, osteoarthritis^17^, and cancer. ^18, 19^ Trace sulfated N-glycans from IgG have also been implicated as potential biomarkers for rheumatoid arthritis. ^20^ A recent study by Zhang *et al*. showed that the cell surface 6-sulfo sialyl LewisX is closely linked to BC metastasis, and could serve as a potential novel marker for BC metastasis. ^21^ These results underscore the importance of sulfated glycans as biomarker candidates and highlight the significance of analyzing sulfated glycans for discovering new biomarkers and elucidating the disease mechanism of BC. ^22, 23^

Despite their intriguing physiological roles, the analysis of sulfated glycans in various mammalian cells, tissues, or organisms faces challenges due to their low abundance. ^24^ Although mass spectrometry (MS)-based analysis is a robust method for glycomic profiling, the analysis of sulfated glycans and other anionic glycans remains challenging. This is primarily due to ion suppression caused by neutral and sialylated non-sulfated glycans in positive^23^ and negative ion modes, respectively. ^24, 25^ Moreover, the negatively charged nature of sulfated glycans, coupled with the presence of labile sulfate groups, poses challenges to their ionization and detection. ^26^ A specialized technique, such as permethylation followed by anion exchange separation^23, 26^, allows for selective detection of sulfated glycans in negative mode, overcoming challenges in separating them from non-sulfated and sialylated glycans. ^23, 25–27^ However, these methods involve multiple cleanup steps, which results in sample loss. ^24^ Recent alternative approaches, such as the use of a serotonin-immobilized column, are effective in analyzing labeled minor acidic glycans, but do not provide information on glycan sialylation status due to prior neuraminidase digestion. ^28^

To address these limitations, we employed a glycoblotting-based sulphoglycomis workflow. The application of this method allows for comprehensive analysis of sulfated and phosphorylated glycans from biological samples. It is particularly effective in enriching benzyloxyamine (BOA)-labeled minor acidic glycans, while simultaneously separating sulfated and phosphorylated glycans^29, 30^, which presents a challenge when using conventional methods due to their similar physicochemical properties and molecular weights (-H_2_PO_3_ = 79.9663 and -HSO_3_ = 79.9568 Da). ^28, 31^

This study represents the first application of the glycoblotting-based sulphoglycomics workflow to analyze the negatively charged sulfated N-glycans expressed in the serum glycoproteins of patients with BC. A comparison was made between the sulfated glycan profiles observed in healthy human serum and patients with BC. Through our analysis, we identified specific sulfated N-glycan species associated with BC. Identifying these glycans has potential diagnostic and prognostic value in clinical settings as they may aid in the detection and management of BC.

## Result

### Analysis of sulfated N-glycans using glycoblotting-based sulphoglycomics workflow

Previously developed methods for the glycoblotting-based sulphoglycomics workflow ^30^ (Supplementary Figure S1) were employed to analyze sulfated N-glycans from human serum samples. This approach enables the integration of glycoblotting-based on-bead glycan enrichment, weak acid group-selective methylation, and labeling on a single platform, coupled with weak anion exchange (WAX) separation and matrix-assisted laser desorption/ionization time-of-flight (MALDI-TOF) MS. First, the glycoproteins in the serum samples were digested with PNGase F to release the N-glycans. Subsequently, glycoblotting, a chemically selective enrichment technique, was used to specifically isolate N-glycans. This offers a straightforward and streamlined approach to enhance the detection and analysis of sulfated N-glycans. Notably, the glycoblotting method demonstrated a higher glycan recovery efficiency from biological samples. ^30^ This means glycoblotting is more effective in separating and enriching sulfated N-glycans from the sample. To eliminate any potential interference during subsequent WAX separation and MALDI-TOF MS, glycoblotting utilizes 3-methyl-1-*p*-tolyltriazene (MTT) for the methyl esterification of the carboxylic group of sialic acid, whereas the sulfated glycans remain unmethylated. ^27, 32, 33^ Therefore, only sulfated glycans retain their negative charges, enabling selective subsequent enrichment using WAX separation and detection in negative mode. WAX separation was used for selective enrichment of sulfated glycans.

This method employs a 3-aminopropyl silica gel microcolumn, which exhibits characteristics similar to those of weak anion-exchange separation methods. ^26^ At neutral pH, the primary amine groups in silica gel act as proton acceptors, resulting in a positive charge. This property is crucial for the retention of negatively charged sulfated glycans. Additionally, the sialic acid groups of the glycans were neutralized in advance by methyl esterification with MTT during glycoblotting. This facilitates the separation of sulfated glycans from the more abundant sialylated glycans, which is typically challenging when analyzing low-abundance sulfated glycans using MALDI-TOF MS in negative mode. Before MS analysis, using WAX separation method, neutral and sialylated N-glycans were initially eluted with a solution of 1% AcOH in 50% MeOH. Subsequently, sulfated N-glycans were eluted using a solution of 1% NH_4_OH in 5% MeOH (pH 10.5).

### MALDI-TOF MS analysis of sulfated N-glycans from healthy human serum samples

As an initial step, MALDI-TOF MS analysis of sulfated N-glycans from healthy human serum samples was performed in negative mode using the glycoblotting-based sulphoglycomics approach. Based on these results, we successfully separated trace sulfated N-glycans from abundant non-sulfated N-glycans (Figure 1). Before enrichment, all non-sulfated N-glycans, including sialylated N-glycans, present in the serum samples were observed, whereas sulfated N-glycans were not identified (Figure 1A). After enrichment, thirteen sulfated N-glycans were detected in the tested samples as [M-H]^-^ ions (Figure 1B). This result clearly shows that MALDI-MS suppressed sulfated glycans in the presence of abundant non-sulfated N-glycans. Among the sulfated N-glycans, in addition to mono-sulfated N-glycans, several di-sulfated N-glycans have also been identified. These glycans are known to significantly enhance binding interactions with numerous glycan-binding proteins, thereby promoting cancer cell migration and invasion. ^34^ No phosphorylated N-glycans were detected.

**Figure 1.**
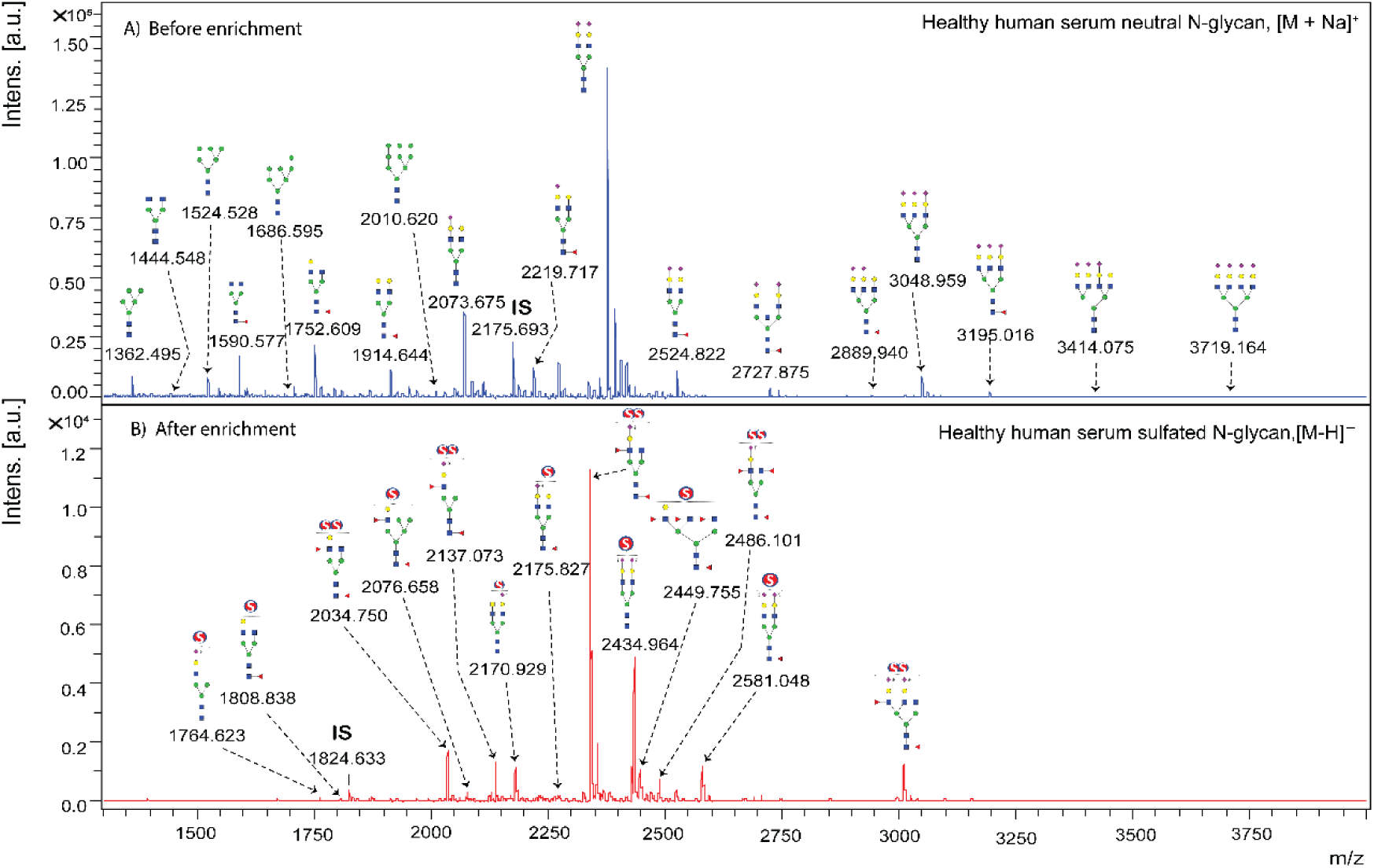
MALDI-TOF MS profiles of BOA-labeled sulfated N-glycans from healthy human serum A) before and B) after enrichment using glycoblotting-based sulphoglycomics workflow. I.S. indicates internal standard.

Compared to non-sulfated N-glycans, the identified sulfated N-glycans are predominantly complex-type structures, with non-reducing terminal sulfo-LacNAc, sulfo-LacdiNAc, sulfo-sialyl-LacNAc, sulfo-lewis-type, and sulfo-sialyl-lewis-type glycan epitopes on N-glycans expressed in human serum samples. Lewis-type epitopes have not been identified for non-sulfated N-glycans (because mono-fucosylated N-glycans are difficult to distinguish from core fucose). In contrast, the major sulfated N-glycans contained multiple fucose residues and presented Lewis-type epitope terminals. Sulfated N-glycans without Lewis-type epitopes also exhibited signal intensity patterns, independent of non-sulfated N-glycans. These findings demonstrate the applicability of the developed method for the analysis of sulfated glycans in serum samples. This method holds promise for identifying new sulfated glycan biomarkers that could be applied in the early detection of BC.

### Quantitative analysis of serum sulfated N-glycans between healthy control individuals and BC patients

We then performed a comparative analysis of the sulfated N-glycan profiles found in the serum samples of 20 normal controls (NC) and 76 BC patients to determine whether these glycans could serve as potential biomarkers for the disease. The clinical information of the study participants is summarized in Supplementary Table S1. Figure 2 shows the representative MALDI-TOF MS profiles of BOA-labeled sulfated N-glycans from BC patients and healthy NC serum samples enriched using the glycoblotting method and then fractioned using WAX. Eight mono-sulfated and five di-sulfated N-glycans were detected in all study samples (Supplementary Table S2). These glycans were selected after each peak was evaluated for their quantitative reproducibility using serially diluted standard serum samples (0.2x, 0.4x, 0.6x, 0.8x, 1x, and 1.2x). The peak area of each glycan was normalized to the concentration of the internal standard, and a standard calibration curve was plotted for each glycan (Supplementary Figure S2). Accordingly, glycan peaks with good slope linearity and minimal outlier scores were selected and considered for further statistical analysis.

**Figure 2.**
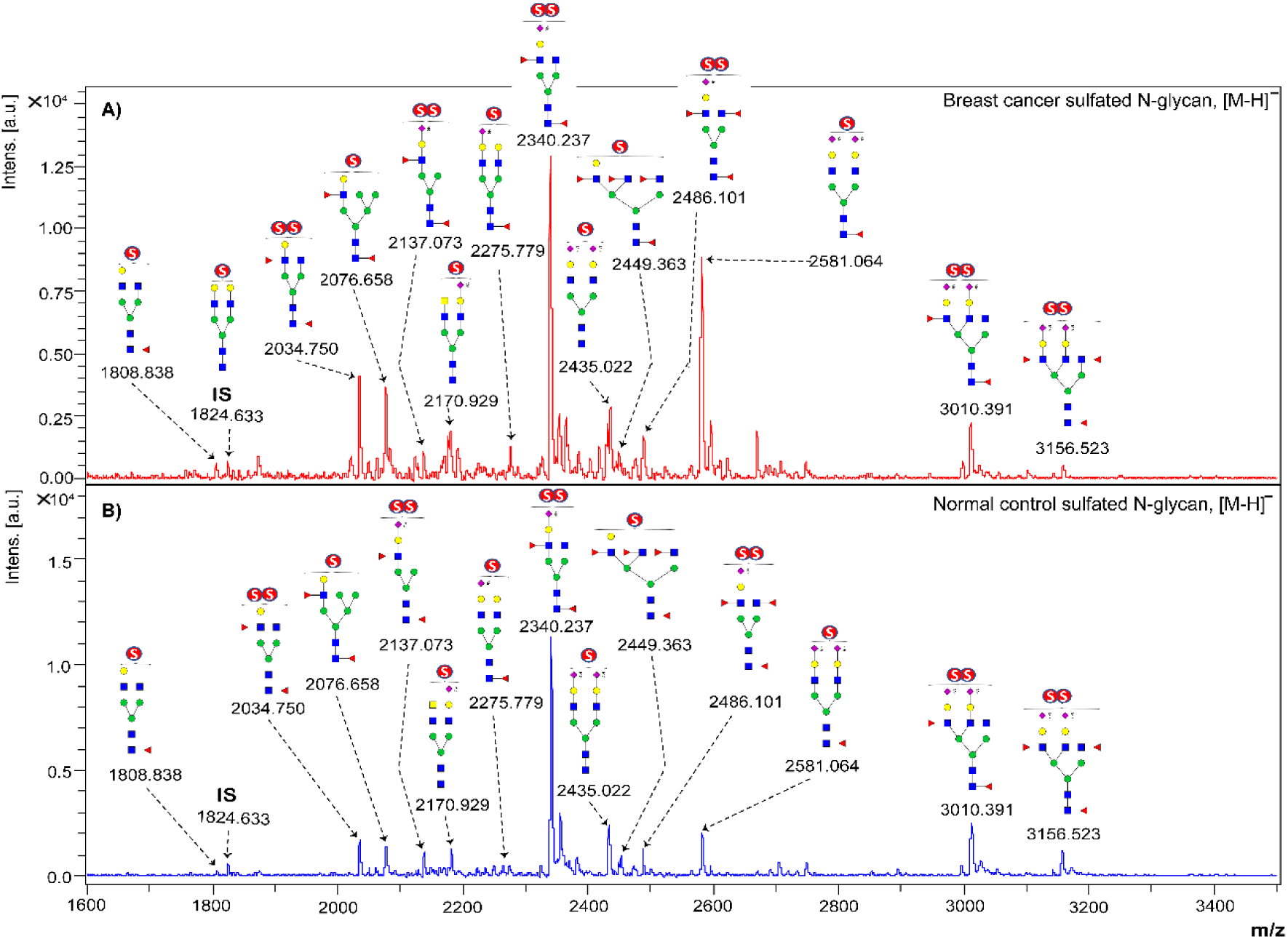
Representative MALDI-TOF MS spectra of BOA-labeled sulfated N-glycans from A) serum BC patients and B) healthy controls. I.S. indicates the internal standard mono-sulfated biantennary complex-type N-glycan (129 pmol).

An internal standard was used to quantitatively analyze the identified sulfated N-glycans in both patients and controls. The internal standard used was mono-sulfated desialylated biantennary complex-type N-glycan (129 pmol), which was synthesized using chemoenzymatic methods from a sialoglycopeptide (SGP). ^35^ A detailed list of all the sulfated N-glycans detected is presented in Supplementary Table S2, along with their corresponding glycan compositions and structures. The results showed that the sulfated N-glycans present in the human serum sample were predominantly complex type, and approximately 1% of the N-glycans were hybrid type. Among complex-type N-glycans, bi-antennary N-glycans comprised the highest proportion (96%), tri- and tera-antennary each comprised 2% and 1%, respectively, and most were fucosylated (≥ 75%) and sialylated (≥ 66%). Comparative analysis revealed a remarkable change in the concentration of individual sulfated N-glycans in human serum samples.

The expression levels of the identified sulfated glycans were determined by calculating the peak area of each glycan after normalization to the known concentration of the internal standard used, as shown in Figure 3. Subsequently, these values were compared between NC and BC groups. An independent t-test was used to identify glycans with differential expression patterns between the groups. The results revealed that seven mono-sulfated and two di-sulfated N-glycans were significantly upregulated in the sera of BC patients compared to the control group, as shown in Table 1. All peaks corresponding to sulfated glycans exhibited a notable increase in abundance (*p* ≤ 0.05) in the sera of BC patients compared with the NC group.

**Figure 3.**
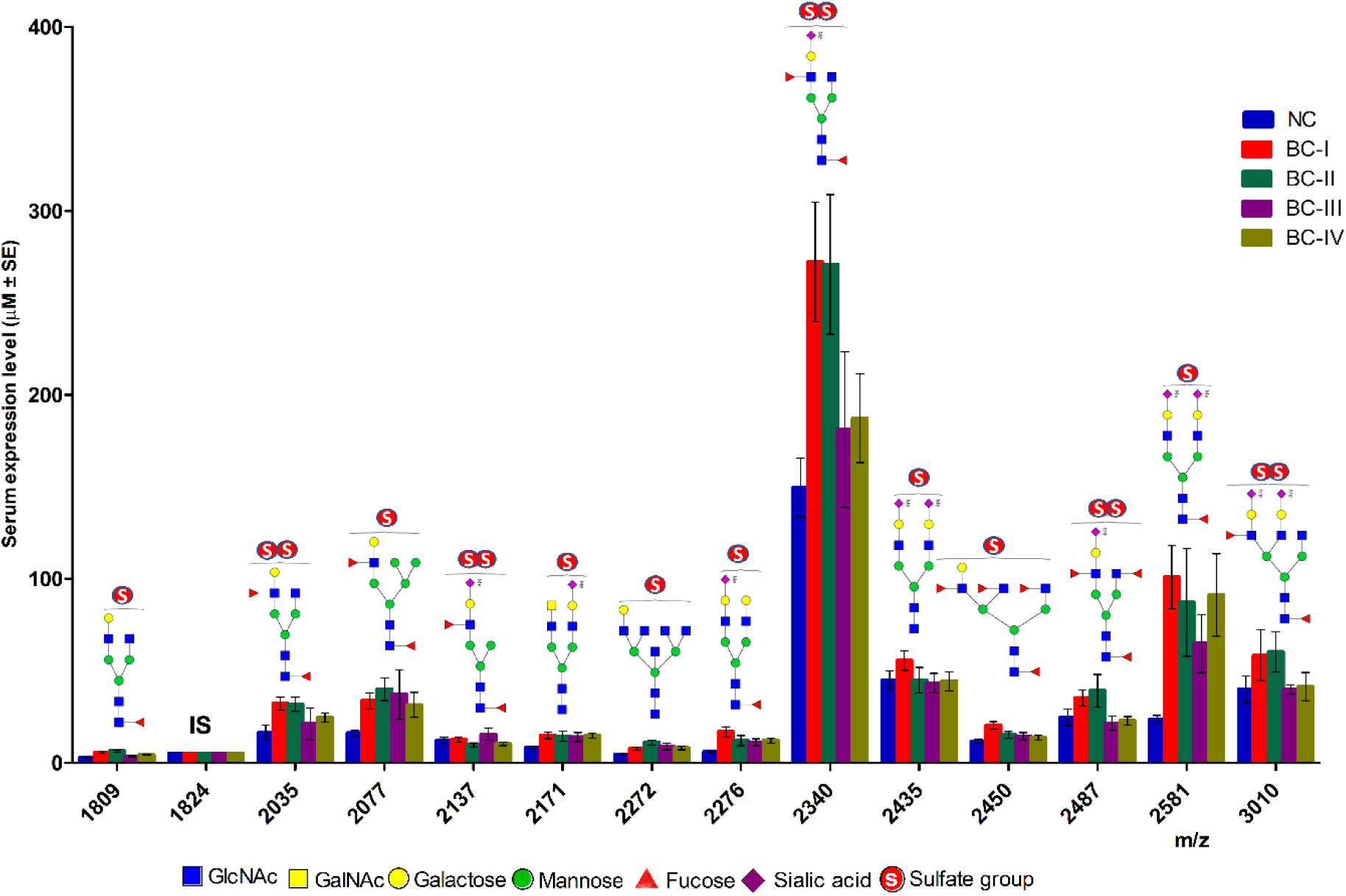
Expression levels and structures of total sulfated N-glycans isolated from NC and BC serum glycoproteins. I.S. indicates an internal standard, mono-sulfated biantennary complex-type N-glycan (129 pmol).

**Table 1.**
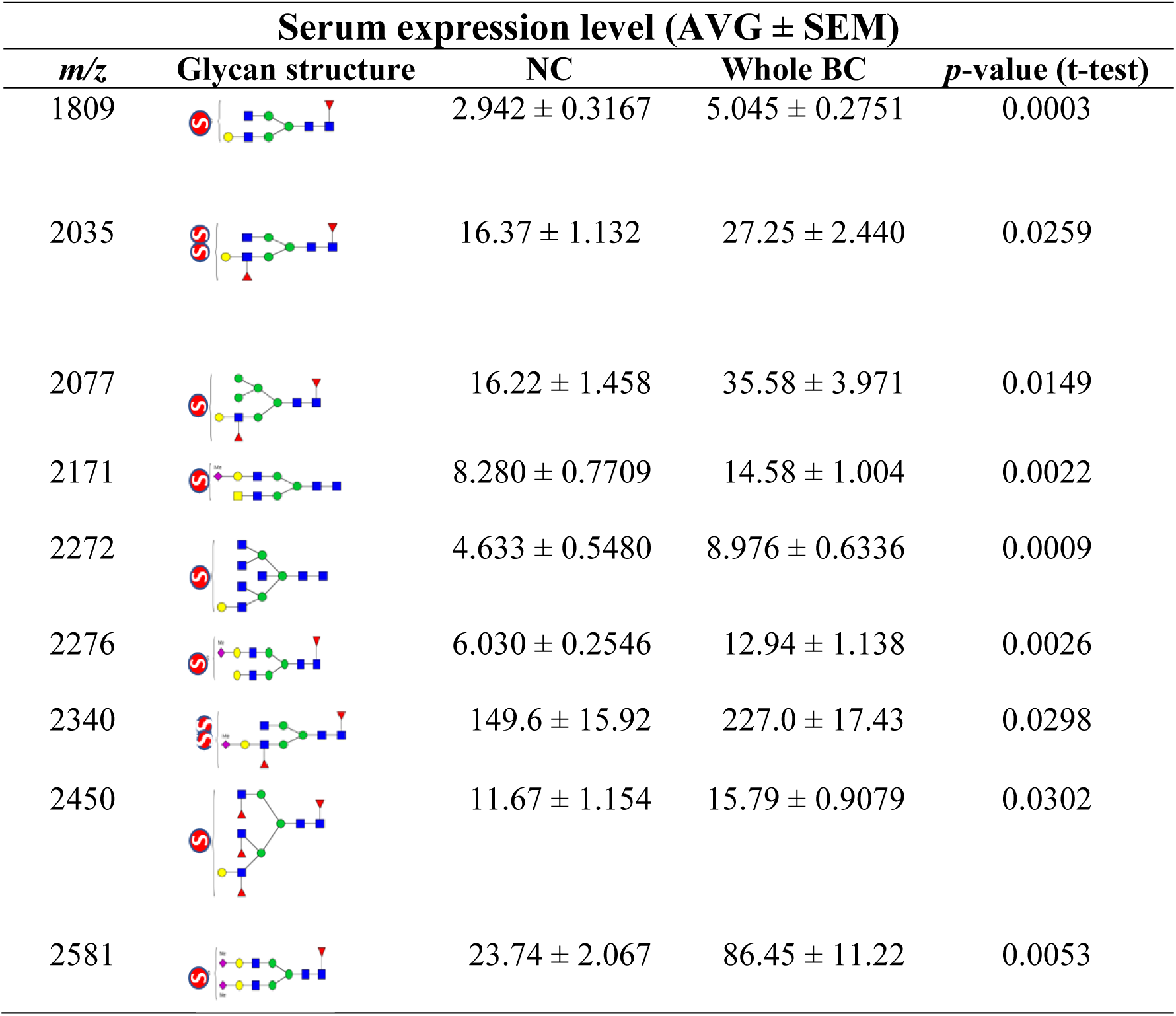
List of upregulated tyssulfated N-glycans in the serum of patients with breast cancer compared with healthy normal controls.

| Serum expression level (AVG ± SEM) |  |  |  |  |
| --- | --- | --- | --- | --- |
| *m/z* | Glycan structure | NC | Whole BC | *p*-value (t-test) |
| 1809 |  | 2.942 ± 0.3167 | 5.045 ± 0.2751 | 0.0003 |
| 2035 |  | 16.37 ± 1.132 | 27.25 ± 2.440 | 0.0259 |
| 2077 |  | 16.22 ± 1.458 | 35.58 ± 3.971 | 0.0149 |
| 2171 |  | 8.280 ± 0.7709 | 14.58 ± 1.004 | 0.0022 |
| 2272 |  | 4.633 ± 0.5480 | 8.976 ± 0.6336 | 0.0009 |
| 2276 |  | 6.030 ± 0.2546 | 12.94 ± 1.138 | 0.0026 |
| 2340 |  | 149.6 ± 15.92 | 227.0 ± 17.43 | 0.0298 |
| 2450 |  | 11.67 ± 1.154 | 15.79 ± 0.9079 | 0.0302 |
| 2581 |  | 23.74 ± 2.067 | 86.45 ± 11.22 | 0.0053 |

### Evaluation of the diagnostic performance of serum sulfated N-glycans

To further examine the diagnostic performance of serum sulfated N-glycans, receiver operating characteristic (ROC) test analysis was employed for glycans that showed statistically significant (*p* ≤ 0.05) changes in expression through t-test analysis. The area under the curve (AUC) values generated from the ROC test were used to evaluate the discriminative potential to differentiate between the cancer patients and control groups. The discriminating potential of candidate biomarkers that met both criteria (*p* ˂ 0.05, AUC ≥ 0.8) was considered a candidate biomarker. Consequently, seven serum sulfated N-glycans showed remarkable diagnostic performance and were considered as candidate biomarkers (Table 2). Notably, these glycan structures are mono-sulfated N-glycans. All seven sulfated glycans were strong predictors to distinguish patients with stage I disease from those in the NC group. These candidates were mono-sulfated LacNAc (*m/z* 1809, 2077, 2272, and 2450), LacdiNAc (*m/z* 2171), and mono- and bi-sialylated (*m/z* 2276, 2581)-terminated N-glycans. Three sulfated glycans (*m/z* 1809, 2077, and 2272) distinguished the stage II patients from the NC group. No significant candidate biomarker was observed for stage III BC patients, whereas four glycans (*m/z* 2077, 2171, 2276, and 2581) fulfilled the criteria for being considered as biomarkers for stage IV patients. Moreover, three sulfated N-glycans (*m/z* 2077, 2276, and 2581) demonstrated the same ability to discriminate all BC patients from the NC group.

**Table 2.** Candidate sulfated N-glycan biomarkers for different BC stages based on ROC analysis.

| *m/z* | Breast cancer VS Normal control |  |  |  |  |  |  |  |  |  | Possible biomarker for |
| --- | --- | --- | --- | --- | --- | --- | --- | --- | --- | --- | --- |
|  | BC-I<br>(n=17) |  | BC-II<br>(n=20) |  | BC-III<br>(n=17) |  | BC-IV<br>(n=22) |  | Whole BC<br>(n=76) |  |  |
|  | AUC | *p* | AUC | *p* | AUC | *p* | AUC | *p* | AUC | *p* |  |
| 1809 | **0.832** | **0.001** | **0.918** | **< 0.0001** | 0.597 | 0.314 | 0.745 | 0.006 | 0.772 | 0.0001 | **I,II** |
| 2077 | **0.832** | **0.001** | **0.857** | **0.0001** | 0.741 | 0.012 | **0.809** | **0.0006** | **0.810** | **< 0.0001** | **I,II, IV, whole BC** |
| 2170 | **0.896** | **0.0001** | 0.726 | 0.015 | 0.685 | 0.054 | **0.838** | **0.0001** | 0.784 | 0.0001 | **I, IV** |
| 2272 | **0.803** | **0.003** | **0.855** | **0.0001** | 0.744 | 0.011 | 0.750 | 0.005 | 0.786 | < 0.0001 | **I, II** |
| 2276 | **0.90** | **< 0.0001** | 0.728 | 0.014 | 0.697 | 0.041 | **0.914** | **< 0.0001** | **0.810** | **< 0.0001** | **I, IV, whole BC** |
| 2450 | **0.803** | **0.0029** | 0.655 | 0.097 | 0.600 | 0.300 | 0.593 | 0.3019 | 0.652 | 0.0382 | **I** |
| 2581 | **0.942** | **< 0.0001** | 0.792 | 0.0018 | 0.708 | 0.030 | **0.918** | **< 0.0001** | **0.840** | **< 0.0001** | **I, IV, whole BC** |

AUC value $\geq 0.8$ and level of significance, $p \leq 0.05$ at 95% CI were considered as criteria
for being the candidate biomarkers (AUC value vs diagnostic accuracy: 0.9-1= highly
accurate, 0.8-0.9= accurate, 0.7-0.8= moderately accurate, $< 0.7$ = uninformative test. The
bold AUC and *p-values* met the criteria and were selected as candidate biomarkers with
their respective stages.

Multiple comparisons were conducted to visualize the abundance of sulfated N-glycans expressed among the different BC stages and the NC group. Figure 4 shows the dot plot and ROC curves for the identified candidate biomarkers related to stage I and stage II BC patients compared with the NC group. The results revealed that a significant increase in glycan abundance was observed primarily during the early stages of the disease.

**Figure 4.**
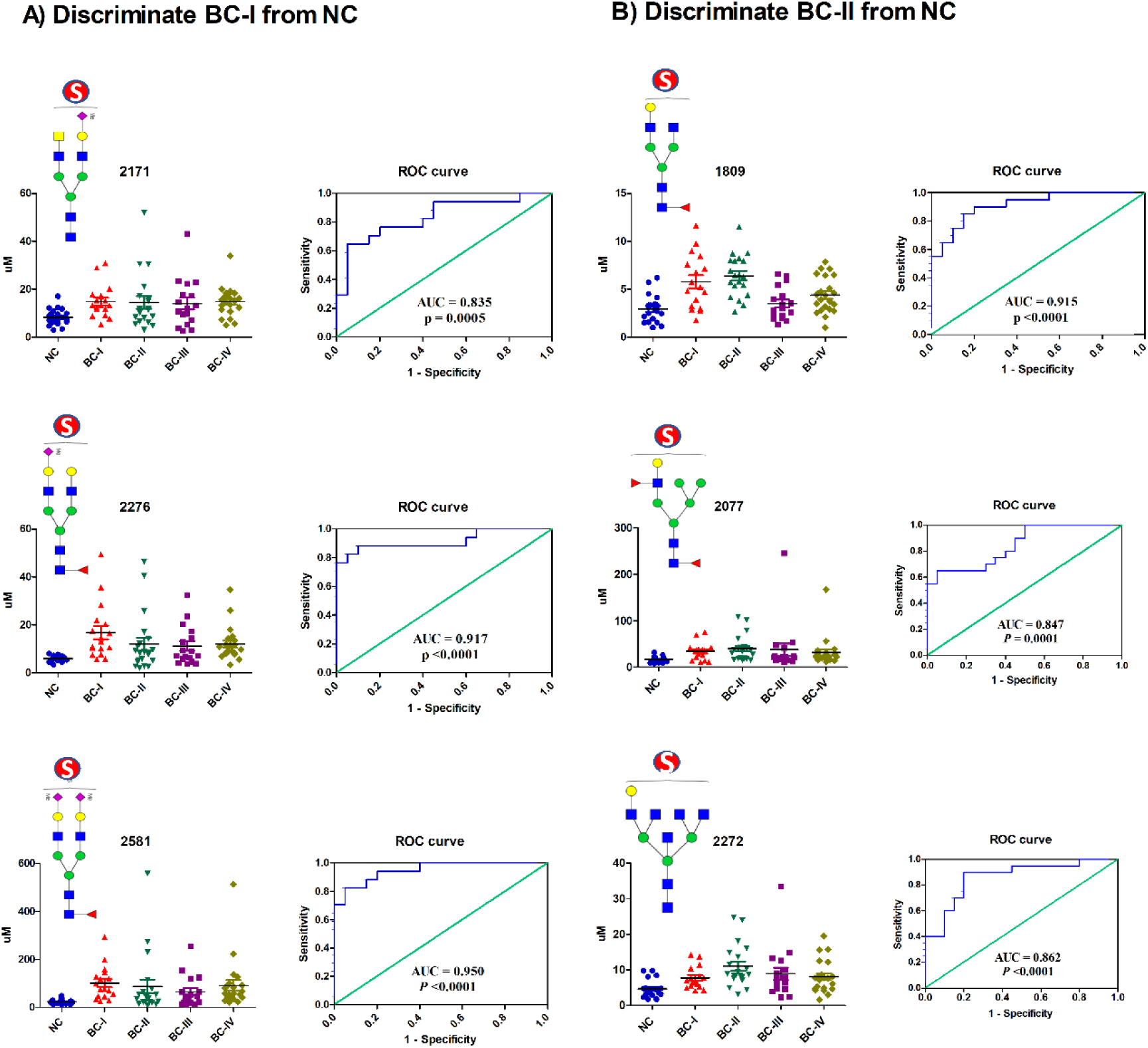
The dot plot illustrates the expression levels of serum sulfated N-glycans that were upregulated in BC patients compared to healthy NC, whereas the ROC curve displays the discriminatory power of each glycan in distinguishing between patients and NC. The area under the curve (AUC) value indicates the effectiveness of glycan as a diagnostic marker: A) Discriminate BC-I from NC; B) Discriminate BC-II from NC.

### Serum glyco-subclass patterns as predictive indicators for early-stage BC

In addition to assessing and comparing the levels of individual glycans between the BC patients and NC groups, we assessed the patterns of serum glycan subclasses. Specifically, we quantified and compared the levels of serum sulfated N-glycans based on their structural similarities between the healthy and cancer groups. Our findings revealed statistically significant variations in the abundance of glycan subtypes, such as fucosylated, bi-antennary, tri-antennary, tetra-antennary, mono-, bi-, and total-sialylated, and total-sulfated N-glycans, in the serum of patients with stage I and II disease (Figure 5). Based on ROC analysis, fucosylated, bi-antennary, mono- and total-sialylated, and total-sulfated were diagnostic indicators of stage I patients. These glycan subtypes demonstrated high AUC values of 0.979, 0.932, 0.944, 0.892, and 0.941, respectively (*p* ˂ 0.0001). In stage II patients, fucosylated and tetra-antennary glyco-subclasses had higher AUC values (0.945 and 0.862, respectively). The increased abundance of these specific glycotypes in the early stages of the disease aligns with the expression patterns of individual sulfated N-glycan biomarkers.

**Figure 5.**
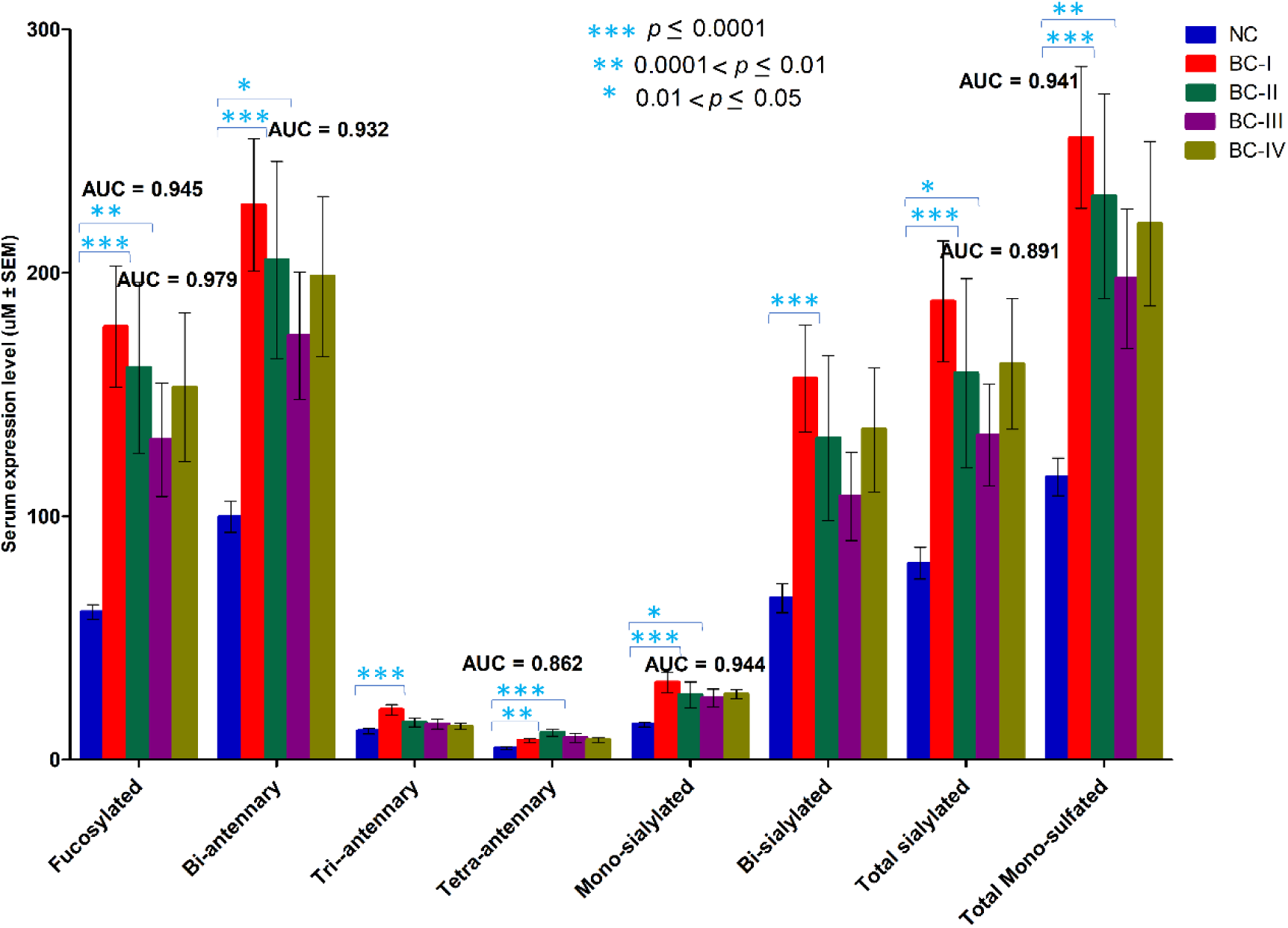
The serum glycan expression patterns of glyco-subclasses shared similar structures between the BC stages and NC group. The area under the curve (AUC) value was used to indicate the effectiveness of glycan subclasses as diagnostic markers for early stage BC.

## Discussion

A growing body of evidence points to the importance of the sulfation of non-reducing terminal epitopes of N-glycans. This sulfation process alters the physicochemical properties of glycans and affects their ability to serve as binding sites for pathogens and endogenous glycan-binding proteins. ^27, 36^ Any alterations in the structure and quantity of sulfated glycans can potentially trigger substantial physiological or pathological processes. ^20^ These terminal sulfate modifications represent carbohydrate antigens associated with tumor malignancy and play pivotal roles in cancer progression and metastasis. ^34, 37^ Therefore, analysis of sulfated N-glycans is of great interest in discovering new biomarkers and elucidating BC disease mechanisms.

In this study, we adapted a glycoblotting-based sulphoglycomics workflow to analyze serum sulfated N-glycans in Ethiopian patients with BC and matched healthy controls. We successfully identified and quantified several mono-sulfated and di-sulfated N-glycans in the serum of patients with BC, which, to the best of our knowledge, have not been previously reported. This indicated the high glycan-capturing efficiency of the glycoblotting method and the high sulfated N-glycan enrichment capacity of WAX separation. We combined this workflow with a synthetic internal standard to achieve efficient quantitative analysis of sulfated N-glycans rich in sialic acid and Lewis-type antigens in serum. This is a noteworthy discovery because it expands our understanding of the glycomic landscape associated with BC.

Differential expression analysis revealed that seven mono-sulfated and two di-sulfated N-glycans were significantly upregulated in the sera of BC patients compared with the control group. ROC analysis revealed that only seven identified mono-sulfated N-glycans exhibited excellent diagnostic performance in distinguishing early stage BC patients from healthy controls (Table 2, Figure S3). These N-glycans had mono-sulfated LacNAc (*m/z* 1809, 2077, 2272, and 2450), LacdiNAc (*m/z* 2171), and mono- and bi-sialylated (*m/z* 2276, 2581) terminals. Early diagnosis is critical for improving patient outcomes and enhancing the accuracy of BC detection, potentially leading to earlier interventions and improved survival rates. Furthermore, based on structural similarity, serum glycotyping patterns exhibited excellent diagnostic performance in specific glycan subtypes for predicting early stage BC. Notably, for stage I, the glycotyping patterns, such as fucosylated, bi-antennary, mono- and total-sialylated, and total-sulfated, demonstrated strong predictive capabilities. At stage II, fucosylated and tetra-antennary glyco-subclasses were robust predictors. These findings suggest noticeable alterations in the biosynthetic pathways of sulfated glycans in BC patients. ^28^ The observed alterations suggest the underlying molecular processes associated with the development and progression of the disease. Investigating these pathways could provide further insights into the biology of BC and potentially reveal novel therapeutic targets.

Furthermore, the significance of our findings is underscored by the discovery that two mono-sulfated N-glycans (*m/z* 2276 and 2581) were identified in human IgG serum samples, as reported by Wang *et al*. ^20^ Moreover, similar to our results, the mono-sialylated complex-type N-glycan (*m/z* 2276) has been recognized as a potential biomarker for rheumatoid arthritis. This was further evidenced by the fact that these two sulfated glycans, along with *m/z* 1809, were previously reported by our groups in their neutral (non-sulfated) forms of human IgG within the same samples. ^12^ This indicated that the three identified biantennary sulfated N-glycans may have originated from serum IgG. These findings strongly suggest an intricate connection between the immune system and alterations in serum sulfate N-glycosylation patterns during BC progression.

Given that IgG is a major serum protein and an essential component of the humoral immune system, N-glycans with sulfation modifications play a crucial role in immune recognition^38^ and can significantly impact the function of IgG, potentially by altering the structure of the Cγ2 domain. ^20^ However, further studies are needed to investigate the effect of sulfated N-glycans on IgG and their association with BC progression. In addition, apart from IgG, carriers of other candidate sulfated biomarkers in human serum have yet to be identified.

We utilized the same serum samples previously examined by Gebrehiwot *et al*. in their study on Ethiopian patients with BC. ^12^ While their analysis focused on identifying non-sulfated N-glycans as potential biomarkers for early BC detection, our investigation revealed a distinct set of sulfated N-glycans. Our findings included specific sulfo-glycotopes that were not identified in their study, indicating a complementary and enriched perspective of the glycomic landscape associated with BC. Their analysis identified 17 candidate biomarkers, whereas our study identified seven sulfated N-glycans that met the criteria for biomarker identification. This suggests a low abundance of sulfated N-glycans. However, despite their lower abundance, sulfated N-glycans exhibited excellent diagnostic accuracy for the early detection of BC compared with the control groups (see Table 2). This insight could provide an additional dimension for identifying potential biomarkers for early detection of BC.

Compared to their non-sulfated counterparts, carbohydrate sulfation has demonstrated increased potency as a ligand for glycan-binding proteins such as selectins^13^ and galectins. ^39^ Studies have illustrated and enhanced the binding affinity of siglecs to their sulfated glycan ligands in various cancer cells, including BC, potentially enhancing their immune invasion capabilities. ^34^ Moreover, it has also been shown that the synthesis of cell surface 6-sulfo sialyl LeX has been implicated in facilitating BC migration and invasions. ^21^ These findings strongly suggest that enhanced sulfation of terminal epitopes on N-glycans could serve as a mechanism for promoting BC progression and metastasis. Similarly, our results showed that sulfated glycans predominantly displayed terminal sulfo-LacNAc, sulfo-LacdiNAc, sulfo-sialyl-LacNAc, sulfo-lewis-type, and sulfo-sialyl-lewis-type glycan epitopes on N-glycans that were upregulated in BC. These modifications might play a role in BC progression, metastasis, and immune invasion. Further exploration is needed to elucidate the specific mechanisms and downstream effects of sulfated N-glycans on BC progression and invasion. Moreover, focusing on these N-glycan pathways may provide a promising therapeutic strategy for BC. ^4^ In particular, targeting the sulfotransferase enzyme responsible for synthesizing specific sulfated N-glycans may impede BC progression and metastasis.

## Conclusion

In conclusion, this study represents the first effort to identify and quantify sulfated N-glycans in the serum of patients with BC, providing new insights into the glycomic profile of the disease. The seven identified sulfated N-glycans exhibited excellent diagnostic accuracy (AUC ≥ 0.8) for detecting early stage BC (BC-I and BC-II). These glycans hold great promise as potential biomarkers of BC. These findings have profound implications for enhancing the diagnostic precision and deepening our understanding of the biology of the disease. Further research and validation is necessary to fully explore the significance of these findings in clinical practice. This study paves the way for future studies involving larger and more diverse cohorts to validate these findings and to explore their clinical utility. They can be integrated into existing diagnostic assays or can serve as the foundation for developing new, highly accurate diagnostic tools.

## Materials and methods

### Study population and sample collection

Human serum samples from female BC patients (n=76) at stages I-IV and age-matched healthy individuals (n=20) were collected from Ethiopia during 2015-2016. Stored serum samples were used in this study. Informed consent was obtained from all study participants and the study was conducted in accordance with the ethical standards outlined in the Declaration of Helsinki. The ethical review boards of Addis Ababa University, School of Medicine, Ethiopia, and Hokkaido University, Faculty of Advanced Life Sciences, Japan approved the research protocols. To ensure sample integrity, serum samples were frozen in plastic vials at −80 °C and carefully packed with dried CO_2_ in a foam box. Subsequently, the serum samples were transported by airplane to Japan within 72 hours for further analysis.

### Materials

Peptide N-glycosidase F (PNGase F) was obtained from New England BioLabs (Ipswich, MA, USA). Ammonium bicarbonate (ABC), 3-methyl-1-p-tolyltriazene (MTT), 2,5-hydroxybenzoic acid (DHB), benzyloxyamine hydrochloride (BOA-HCl), sodium bicarbonate (NaHCO_3_), trifluoroacetic acid (TFA), and 3,4-diamino benzophenone (DABP) were purchased from Tokyo Chemical Industry Co. (Tokyo, Japan). BlotGlyco H beads were purchased from Sumitomo Bakelite, Co. Ltd. (Tokyo, Japan), proteinase K was from Roch (Germany), and trypsin was from Sigma-Aldrich Corp. (St. Louis, MO, USA). MultiScreen Slvinert^R^ filter plates were purchased from Millipore Co., Inc. (Tokyo, Japan).

### N-Glycan release from Human Serum Glycoprotein ^30, 40^

Whole serum from each sample was subjected to enzymatic pretreatment to generate glycans with a reducing terminal. This enzymatic pretreatment facilitated subsequent chemical ligation with hydrazide-functionalized BlotGlyco^®^ H beads during the glycoblotting process. Hence, 15 µL of human serum and 10 µL of a 12.9 µM mono-sulfated desialylated N-glycan prepared from SGP (Tokyo Chemical Industry Co., Ltd.), used as an internal standard, were mixed with 50 µL of 200 mM NH_4_HCO_3_. 4 µL of denaturing buffer (5% sodium dodecyl sulfate, 0.4 M dithiothlotol) was added to denature the sample, followed by heating at 100 °C for 10 minutes. Subsequently, 10 µL of 123 mM iodoacetamide (IAA) was added and incubated at room temperature in the dark for 1 hour. The mixture was then subjected to digestion by adding 10 µL of trypsin (40 U/ µL) dissolved in 1 mM hydrochloric acid (HCl) and incubated overnight at 37 °C. Afterward, enzyme activity was halted by heating at 90 °C for 10 minutes and allowed to cool to room temperature. To initiate further processing, 8 µL of reaction buffer (0.5 M Na_3_PO_4_, pH 7.5) and 10% NP-40 were sequentially added and incubated for 10 minutes at 37 °C. N-Glycans were released using 2 µL of PNGase F (New England BioLabs) with an activity of 5 U/μL, followed by overnight incubation at 37 °C. For further digestion,10 µL proteinase K (0.5 U/ µL) was added and incubated for 3 hours at 37 °C. The enzyme was then heat inactivated at 90 °C for 10 minutes. The resulting sample was dried using SpeedVac and stored at −20 °C until further use.

### Enrichment of glycans using glycoblotting^30, 40–42^

A 250 µL suspension of BlotGlyco^®^ H beads (Sumitomo Bakelite Co., Ltd.) with a concentration of 10 mg/mL in water was aliquoted into 96 wells of a MultiScreen Solvinert filter plate (Millipore, Billerica, MA, USA), and the water was removed by applying vacuum. The dried sample containing released N-glycans was reconstituted by adding 20 µL of Milli-Q water. The reconstituted sample (20 µL) was added to each well, followed by 180 µL of 2% acetic acid in acetonitrile (AcOH/ACN). The plate was incubated at 80 °C for 45 minutes. Following the incubation, two successive washing steps were carried out using 200 µL each of a 2 M guanidine-HCl in 16.6 mM NH_4_HCO_3_, water, and a 1% solution of triethylamine in methanol (MeOH). The unreacted hydrazide functional groups were acetyl-capped by adding 100 µL 10% acetic anhydride in MeOH at room temperature for 30 minutes. Any residual acetic anhydride was removed under a vacuum. Each well was washed twice with 10 mM HCl, MeOH, and dioxane. For on-bead methyl esterification, 100 µL of 100 mM MTT in dioxane was added and the mixture was incubated at 60 °C for 90 minutes. Subsequently, two washing steps were performed using 200 µL of dioxane, water, MeOH, and water. To facilitate effective labeling through a trans-iminization reaction, 20 µL of 50 mM BOA-HCl and 180 µL of 2% AcOH/ACN solution were sequentially added. The mixture was then incubated at 80 °C for 45 minutes. N-Glycans labeled with BOA were eluted twice with water (150 µL). The resulting glycan solution was dried using SpeedVac and stored at −20 °C until further use.

### Enrichment of sulfated N-glycans using WAX ^29, 30^

Enrichment of sulfated N-glycans was performed according to previously reported methods. ^30^ A small cotton plug was inserted into a 200 microliters micropipette tip. Subsequently, 50 µL 3-aminopropyl silica gel (Tokyo Chemical Industry Co. Ltd., 100 mg/mL suspension) was loaded onto the micropipette tip on top of a cotton plug. The gel was allowed to settle under gravity. The packed WAX microcolumn was conditioned and washed twice with 100 µL water, ACN, and 1% AcOH in 95% ACN. After each washing step, the mixture was centrifuged at 500 rpm for 2 minutes. To reconstitute the BOA-labeled N-glycans obtained from glycobolotting, 20 µL of MilliQ water was used. A 5 µL sample aliquot was then dissolved in 150 µL of 1% AcOH in 95% ACN, which was then loaded into the column. The sample was allowed to elute by gravity. The collected eluate was reloaded into the column, and this process was repeated three times to ensure adequate interaction. The column was washed with 150 μL of 1% AcOH in 95% ACN, followed by centrifugation at 500 rpm for 2 minutes to remove unbound and hydrophobic contaminants. Then, 150 µL of 1% AcOH in 50% ACN was added to elute BOA-labeled neutral and methylated sialylated N-glycans. Elution was performed twice, and after each elution, centrifugation was conducted at 500 rpm for 2 minutes. Sulfated N-glycans were eluted using 150 µL 1% NH_4_OH in 5% ACN (pH 10.5). As in the previous steps, this elution was repeated twice, and centrifugation at 500 rpm for 2 minutes was performed. Finally, the resulting glycan solution was dried using SpeedVac and stored at −20 °C until use.

### MALDI-TOF MS analysis

Mass spectrometric analysis of BOA-labeled sulfated N-glycans was performed using an Ultraflex III (Bruker, Germany) equipped with a reflector in negative ion mode. DABP (10 mg/mL) in 75% ACN with 0.1% TFA was used as a matrix throughout this work^25, 43^, from which 0.5 µL of each sample and matrix solution were spotted onto an MTP 384 target plate (polished steel TF, Bruker Daltonics) and dried at room temperature. To ensure reproducibility, each sample was spotted in four replicates, and each spectrum was generated by accumulating 1000 laser shots. The average of the four normalized data points for each sample was used for statistical analysis. Data acquisition and processing were performed using the Flexanalysis 3.0 software (Bruker, Germany). The intensity of the monoisotopic peak for each sulfated N-glycan was normalized using 12.9 µM of an internal standard (mono-sulfated biantennary complex-type N-glycans synthesized using chemoenzymatic methods^35^). The normalized data were used for further statistical analysis and quantitative comparison of sulfated N-glycan profiles. The structural compositions of sulfated N-glycans were assigned using the Expasy GlycoMod Tool and Glyconnect Database provided by the Swiss Institute of Bioinformatics (https://web.expasy.org/glycomod/), and GlycoWorkbench. ^44, 45^ GraphPad Prism 5 software was used for data analysis and graph plotting.

### Statistical Analysis

A comparison between the two groups (NC vs. entire BC group) was performed using an independent sample t-test. Multiple comparisons among the NC and clinical stage groups were conducted using Bonferroni one-way analysis of variance (ANOVA). To evaluate the diagnostic potential of individual glycans or glyco-subclasses that showed significant differences, a receiver operating characteristic (ROC) test was performed. The area under the curve (AUC) value generated from the ROC test was used as a measure of the diagnostic accuracy of potential glycan biomarkers. AUC values within the ranges of 0.9-1, 0.8-0.9, 0.7-0.8, and <0.7 are classified as “highly accurate,” “accurate,” “moderately accurate,” and “uninformative tests,” respectively. Differences in mean values were considered significant at a 95% confidence interval (*p* ≤ 0.05).

## Data Availability

All data are available in the main text or supplementary materials.

## Authors’ Contribution

**D. G. Feleke:** Data curation, formal analysis, investigation, validation, writing-original draft, writing-review and editing. **B. M. Montalban:** Data curation, methodology, validation, writing-review and editing. **S. T. Gizaw:** Conceptualization, funding acquisition, project administration, resources, validation, writing-review and editing. **H. Hinou:** Conceptualization, data curation, funding acquisition, project administration, methodology, project administration, resources, supervision, validation, writing-original draft, writing-review and editing.

## Acknowledgments

This work was supported by the Japan Society for the Promotion of Science Grant-in-Aid for Scientific Research (22H02191 to H. Hinou), Core-to-Core type B to H. Hinou, Breast Cancer Thematic Research from Addis Ababa University (to S. T. Gizaw). Authors thank the study participants from Tikur Anbesa Specialized Hospital, CHS, Addis Ababa University and Saint Paul’s Millennium Medical College, Addis Ababa, Ethiopia. The authors appreciate to the supports in the laboratory managements by Ms. Maki Morita and Ms. Tomoko Takahashi. B. M. Montalban hanks the MEXT scholarship under the International Graduate Program (IGP), Hokkaido University. D. G. Feleke thanks the MEXT scholarship under the Super Global University (SGU) Program, Hokkaido University.

## Data and material availability

All data are available in the main text or supplementary materials.

