## Supplementary material for "Sulfated N-glycans Upregulation in Sera Predicts Early-Stage Breast Cancer in Patients": Figure S1-S3, Table S1 and S2

### Supplementary information

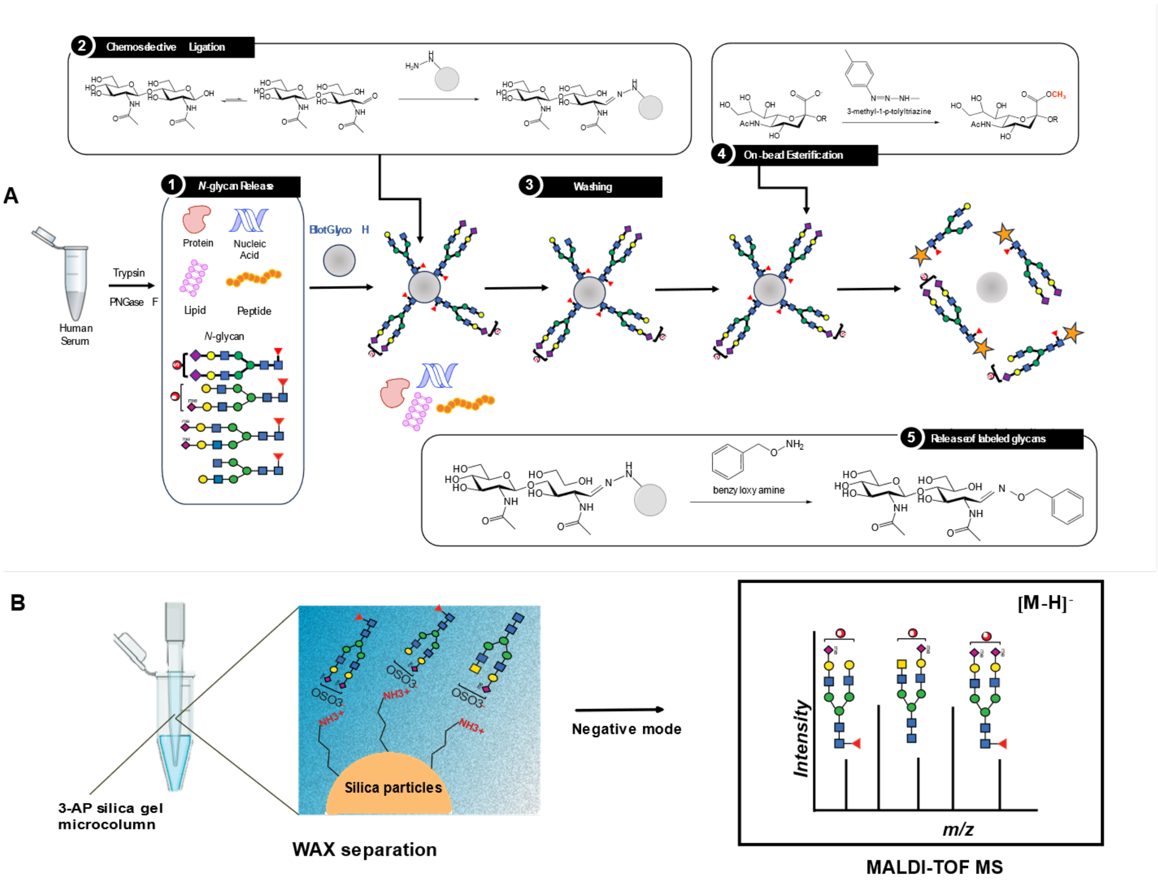


**Figure S1**. Schematic workflow of the glycoblotting-based sulphoglycomic approach for sulfated N-glycan analysis. A) Overall workflow of the glycoblotting-based MALDI-TOF M/S analysis of N-glycans derived from serum glycoproteins. 1) Enzymatic releasing of N-glycans from serum glycoproteins 2) Chemoselective ligations of N-glycans by capturing reducing sugars onto hydrazide-functionalized BlotGlyco H beads 3) Washing to remove impurities 4) On-bead methyl esterification of –COOH of terminal sialic acid residues 5) Recovery of Benzyloxyamine (BOA)-labeled glycans using trans-iminization reaction. B) Weak anion exchange (WAX) separation using a 3-aminopropyl (3-AP) silica gel microcolumn, followed by MALDI-TOF M/S analysis in negative mode.

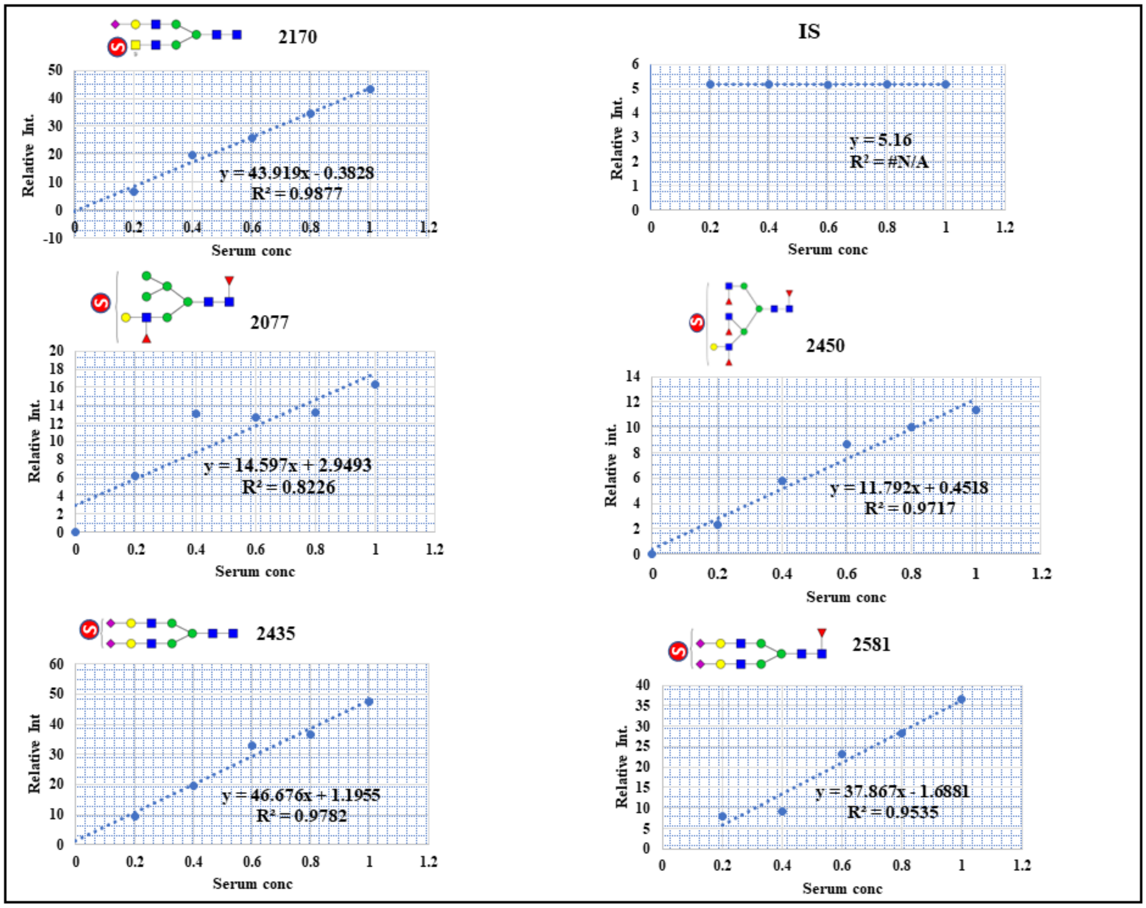


**Figure S2**: Human serum calibration curve showing the quantitative reproducibility of common sulfated glycans.

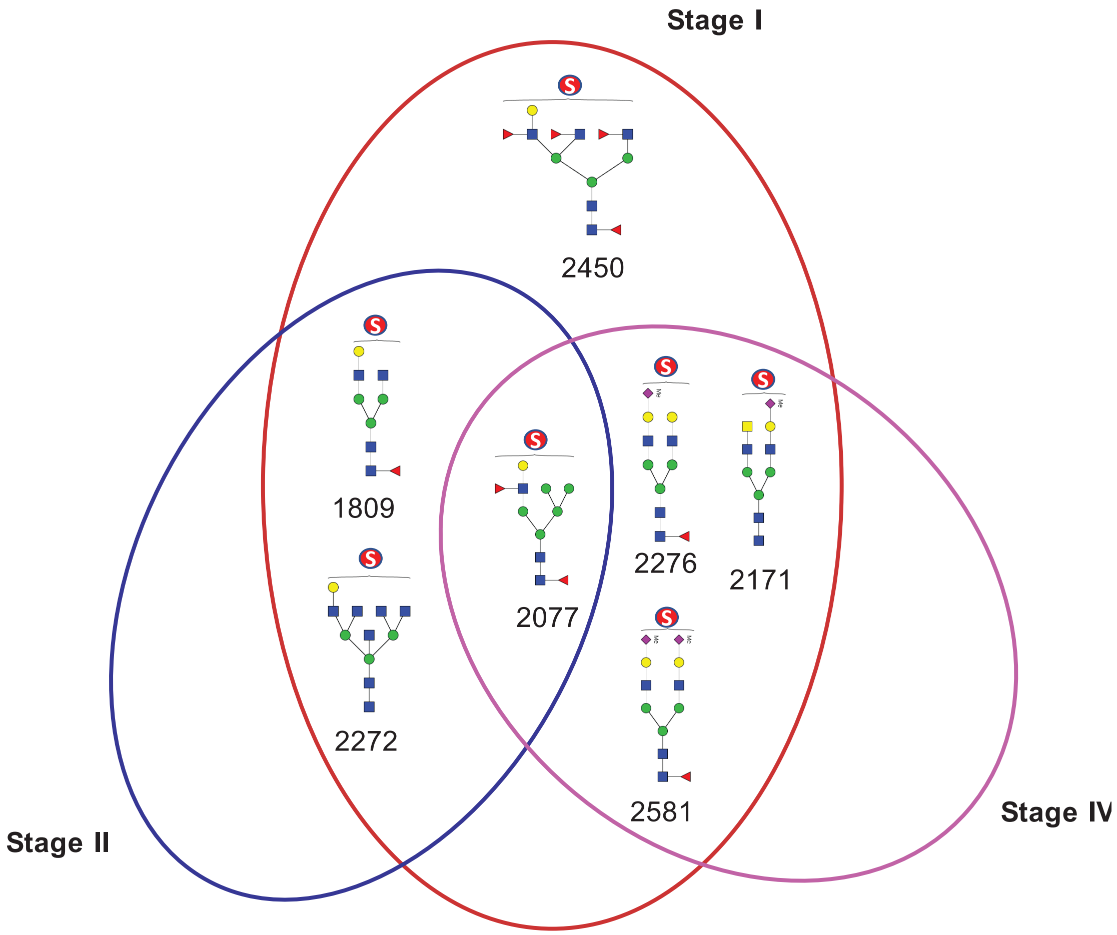


**Figure S3**. Venn diagram illustrating BC stage-specific and overlapping sulfated N-glycans identified as candidate biomarkers.

#### Table S1. Demographic characteristics of the study participants

| | | Mean ± SD | | Number of subjects per age range | | | | |
|---|---|---|---|---|---|---|---|---|
| **Status** | **Number (n)** | **Age** | **BMI (kg/m2)** | **20-30** | **31-40** | **41-50** | **51-60** | **≥61** |
| NC | 20 | 32.3 ± 7.54 | 22.33 ± 3.20 | 10 | 8 | 1 | 1 | |
| BC-I | 17 | 43.0 ± 13.88 | 22.37 ± 2.56 | 4 | 5 | 3 | 3 | 2 |
| BC-II | 20 | 42.8 ± 11.23 | 21.90 ± 3.10 | 3 | 7 | 5 | 2 | 3 |
| BC-III | 17 | 39.7 ± 11.02 | 22.90 ± 2.66 | 3 | 9 | 4 | | 1 |
| BC-IV | 22 | 40.36 ± 12.29 | 22.86 ± 2.87 | 6 | 7 | 6 | 2 | 1 |
| **Total (N)** | **96** | | | **26** | **36** | **19** | **8** | **7** |

**Table S2: Estimated compositions of sulfated N-glycans from human serum glycoproteins labeled with BOA.**

| Observed mass, *m/z* $[M-H]^-$ | Calculated mass, *m/z* $[M-H]^-$ | Error ppm | Glycan composition | Probable structure [1-3] | Glyconnect database links |
|---|---|---|---|---|---|
| 1764.623 | 1764.589 | 25.06 | $(Hex)_1$ $(HexNAc)_1$ $(NeuAc)_1$ $(Sulph)_1$ + $(Man)_3(GlcNAc)_2$ | 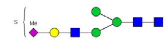 | |
| 1808.838 | 1808.615 | 120.09 | $(Hex)_1$ $(HexNAc)_2$ $(dHex)_1$ $(Sulph)_1$ + $(Man)_3(GlcNAc)_2$ | 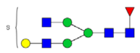 | GlyConnect |
| 1824.629 | 1824.610 | 7.79 | $(Hex)_2$ $(HexNAc)_2$ $(Sulph)_1$ + $(Man)_3(GlcNAc)_2$ | 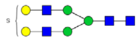 | GlyConnect |
| 1913.718 | 1913.646 | 34.60 | $(Hex)_2$ $(HexNAc)_2$ $(dHex)_2$ $(Sulph)_1$ + $(Man)_3(GlcNAc)_2$ | 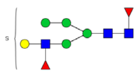 | |
| 2034.750 | 2034.63 | 56.63 | $Hex)_1$ $(HexNAc)_2$ $(dHex)_2$ $(Sulph)_2$ + $(Man)_3(GlcNAc)_2$ | 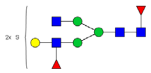 | |
| 2076.558 | 2075.699 | 410.88 | $(Hex)_3$ $(HexNAc)_1$ $(dHex)_2$ $(Sulph)_1$ + $(Man)_3(GlcNAc)_2$ | 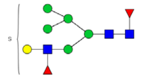 | |

| | | | | | |
|---|---|---|---|---|---|
| 2130.004 | 2129.721 | 137.66 | (Hex)$_2$ (HexNAc)$_2$ (NeuAc)$_1$ (Sulph)$_1$ + (Man)$_3$(GlcNAc)$_2$ | 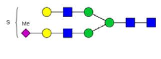 | GlyConnect |
| 2137.073 | 2136.66 | 195.57 | (Hex)$_1$ (HexNAc)$_1$ (dHex)$_2$ (NeuAc)$_1$ (Sulph)$_2$ + (Man)$_3$(GlcNAc)$_2$ | 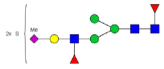 | |
| 2170.980 | 2170.747 | 112.04 | (Hex)$_1$ (HexNAc)$_3$ (NeuAc)$_1$ (Sulph)$_1$ + (Man)$_3$(GlcNAc)$_2$ | 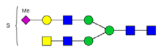 | GlyConnect |
| 2271.621 | 2271.795 | -79.13 | (Hex)$_1$ (HexNAc)$_5$ (Sulph)$_1$ + (Man)$_3$(GlcNAc)$_2$ | 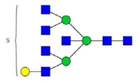 | |
| 2275.807 | 2275.779 | 16.79 | (Hex)$_2$ (HexNAc)$_2$ (dHex)$_1$ (NeuAc)$_1$ (Sulph)$_1$ + (Man)$_3$(GlcNAc)$_2$ | 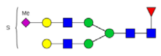 | GlyConnect |
| 2340.237 | 2339.741 | 216.31 | (Hex)$_1$ (HexNAc)$_2$ (dHex)$_2$ (NeuAc)$_1$ (Sulph)$_2$ + (Man)$_3$(GlcNAc)$_2$ | 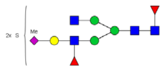 | |
| 2435.022 | 2434.832 | 88.8 | (Hex)$_2$ (HexNAc)$_2$ (NeuAc)$_2$ (Sulph)$_1$ + (Man)$_3$(GlcNAc)$_2$ | 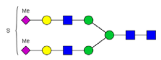 | |
| 2449.363 | 2449.868 | 208.53 | (Hex)$_1$ (HexNAc)$_3$ (dHex)$_4$ (Sulph)$_1$ + (Man)$_3$(GlcNAc)$_2$ | 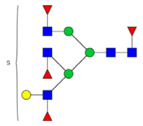 | |
| 2486.101 | 2485.798 | 125.59 | (Hex)$_1$ (HexNAc)$_2$ (dHex)$_3$ (NeuAc)$_1$ (Sulph)$_2$ + (Man)$_3$(GlcNAc)$_2$ | 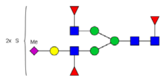 | |
| 2581.064 | 2580.890 | 77.57 | (Hex)$_2$ (HexNAc)$_2$ (dHex)$_1$ (NeuAc)$_2$ (Sulph)$_1$ + (Man)$_3$(GlcNAc)$_2$ | 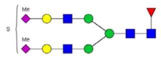 | GlyConnect |
| 3010.391 | 3009.984 | 143.91 | (Hex)$_2$ (HexNAc)$_3$ (dHex)$_2$ (NeuAc)$_2$ (Sulph)$_2$ + (Man)$_3$(GlcNAc)$_2$ | 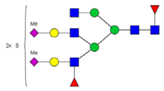 | |
| 3156.523 | 3156.042 | 160.69 | (Hex)$_2$ (HexNAc)$_3$ (dHex)$_3$ (NeuAc)$_2$ (Sulph)$_2$ + (Man)$_3$(GlcNAc)$_2$ | 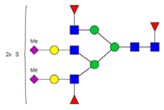 | |

The *m/z* 1824 was the internal standard (IS). The *m/z* values of 1764.623, 1913.718, 2130.004, and 3156.523 were not included in the quantitative comparison because they were not quantitatively reproducible for the entire study sample. Monosaccharide nomenclatures are based on SNFG: Hexose (Hex), *N*-acetyl hexosamine (HexNAc), mannose (Man), fucose (dHex), N-acetyl glucosamine (GlcNAc), sulfate (Sulph), and *N*-acetyl neuraminic acid (NeuAc).
